# The real time effective reproductive number for COVID-19 in the United States

**DOI:** 10.1101/2020.05.08.20095703

**Authors:** Yue Zhang, Lindsay T. Keegan, Yuqing Qiu, Matthew H. Samore

**Affiliations:** University of Utah, Study Design and Biostatistics Center, Salt Lake City, Utah; University of Utah, Department of Internal Medicine, Division of Epidemiology, Salt Lake City, Utah; Department of Veterans Affairs Salt Lake City Health Care System, Salt Lake City, Utah, USA

## Abstract

none.

---

Severe-acute respiratory syndrome coronavirus 2 (SARS-CoV-2), the virus responsible for COVID-19, has spread rapidly causing significant global morbidity and mortality^1^. The United States has now emerged as the global epicenter. As states move to lift intervention measures, it becomes more important to estimate the rate at which the epidemic is growing using the realtime effective reproductive number, R_t_. Using a collated time series of daily state-wise positive case counts from the COVID Tracking Project ^2^, we estimate R_t_ to provide a baseline estimate of the impact of all combined non-pharmaceutical interventions (NPIs) at the state level and for the entire country. We present the results for each state and the whole country in a user-friendly web interface updated daily at https://covid19-realtimereproductionnumber.shinvapps.io/ShinvAPP/. We calculate R_t_ using two complementary methods, the Wallinga and Teunis (WT) method ^3^ which is forward-looking and the Cori method ^4^ which is backward-looking. We also present two daily test metrics: test utilization per capita and the positive tests per capita ^2^ alongside R_t_ for each state (See Figure 1 with Utah and the U.S. as examples). The results for all 50 states and the U.S. are provided as supplementary materials.

**Figure 1:**
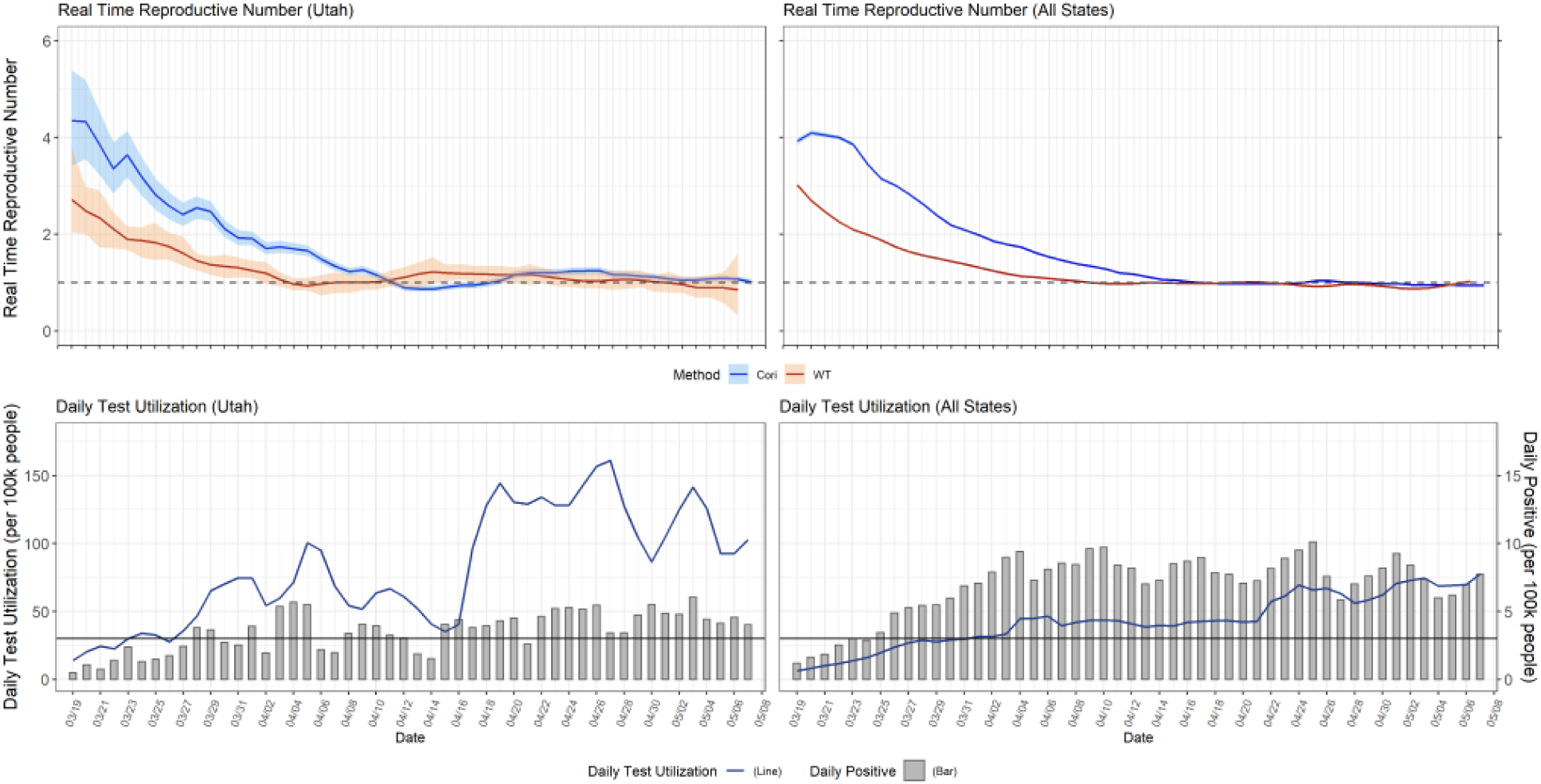
Estimates of the real time effective reproductive number, R_t_, for Utah and the U.S. from March 19 until May 7, 2020. Top Panel: Estimates of R_t_ using two methods, WT (red) and Cori (blue) for Utah (left) and the U.S. (right), respectively. The generation time distribution is assumed to follow Gamma distribution with mean 6.5 and standard deviation 4 for both methods. Bottom Panel: Daily test utilization per 100,000 people (grey bars) and daily positive tests per 100,000 people (blue line) for Utah (left) and the U.S. (right), respectively.

The overall downward trend indicates that most states have been able to reduce the reproductive number of SARS-CoV-2. However, few states have demonstrated an ability to maintain R_t_ below one in a statistically significant manner. An uptick of R_t_ around the beginning of May might indicate that political protests or exhaustion for social distancing may be impacting our ability to control COVID-19. While we find that both methods show a drop in R_t_, we find a significant difference in the timing of R_t_<1. As noted in Lipsitch et al. 2020 ^5^, the difference is consequential enough to impact policy decision making, highlighting the importance of method selection for R_t_ estimation. We find that the median difference in timing of R_t_<1 between the two methods is approximately one week.

Although data quality issues plagued early estimates of R_t_, we believe our estimates of R_t_ are beneficial for tracking the impacts of lifting restrictions on social activity and reducing NPIs. Given the utility of R_t_, we plan to continue hosting and managing the web interface throughout the COVID-19 pandemic. We believe this product is crucial to help policy makers track the progress of current policies and modulate future risk levels.

## Data Availability

The data are already publicly available

